# Association of occupational exposure to chemical substances with bladder cancer in Ethiopia: A multi-center matched case-control Study

**DOI:** 10.1101/2024.09.05.24313148

**Authors:** Eleni Asfaw Kebede, Tigist Workneh Leulseged

**Author notes:** Corresponding author: Eleni Asfaw Kebede Email address (EAK).

## Abstract

**Background:** Bladder cancer (BC) is the 10th most common cancer worldwide and ranks 13th in incidence in Ethiopia. Occupational exposure is the second leading cause of BC after smoking, accounting for 21%–27% of BC cases in men and 11% in women. This study aims to investigate the association between occupational exposure to chemical substances and bladder cancer among patients in selected hospitals in Ethiopia.

**Methods:** A multi-center matched case-control study was conducted among patients admitted to four teaching hospitals in Addis Ababa, Ethiopia from May 01 to August 31 2023. Underlying characteristics between cases and controls were compared using Chi-square test and Mann-Whitney U test with statistical significance set at p ≤ 0.05. A multivariable binary logistic regression analysis was run where adjusted odds ratios (AOR) with 95% confidence intervals (CIs), and p-values were used for significance testing and result interpretation.

**Results:** Among the 303 patients studied, 90 (29.7%) were cases and 213 (70.3%) were controls. Age (±5 years), sex, and smoking status were matched in 68% of participants. The majority of the participants were age 60 years and above (62.2% of cases, 59.1% of controls), male (72% of cases, 73% of controls) and non-smokers (61.1% of cases, 64.8% of controls). Occupational exposure was not found to be significantly associated with BC (AOR:1.09; 95%CI= 0.44, 2.74; P=0.85). On the other hand, having a primary level education or higher (AOR:0.12 95%CI= 0.05, 0.30; P<0.001), consuming coffee for more than 20 years (AOR:2.36; 95% CI=1.22, 4.56; P=0.01) and regular analgesics use (AOR: 6.60; 95% CI=3.34, 13.02; p<0.001) were found to be significantly associated with BC.

**Conclusions:** Occupational exposure to chemical substances was not found to be significantly associated with BC. However, having primary education or higher was protective, while long-term coffee consumption and regular analgesic use were significantly associated with BC. Health care professionals are advised to educate patients about safe analgesic use. Furthermore, future multicenter case-control studies should be conducted to further explore the potential associations of these factors with bladder cancer.

## INTRODUCTION

Bladder cancer (BC) is the 10th most common cancer globally, with an increasing incidence worldwide (1). In 2020, there were over 573,000 new cases and 212,536 deaths due to BC (2). Based on the data from global cancer observatory 2022, in Ethiopia, BC ranked as the 13th most common cancer, with 1,647 new cases and 923 deaths in 2022 (3).

Occupational exposure to chemical substances, which is the second leading cause of BC after smoking, accounts for 21%-27% of cases in men and 11% in women (4, 5). A 2015 meta-analysis identified increased BC incidence in 42 out of 61 occupational classes, particularly among workers exposed to aromatic amines and polycyclic aromatic hydrocarbons (PAHs) (6). While Western countries have experienced a decline in BC incidence attributed to the banning of occupational chemicals, developing countries encounter heightened risks due to urbanization, industrialization, and the transfer of manufacturing processes from developed nations (7). According to the International Labor Organization (ILO), only 5%-10% of workers in developing countries have access to occupational health services (OHS) (8). The lack of stringent occupational safety measures exacerbates the burden of BC in these countries (7).

In addition, BC is associated with several risk factors, including male sex, older age, obesity, and genetic predispositions such as having a first-degree relative with BC and blood group O (5, 6, 9, 10). Environmental factors, such as residence in industrialized areas, pelvic radiotherapy, cyclophosphamide chemotherapy, and chronic bladder infections, also contribute to BC risk (11, 12, 13). Furthermore, Schistosoma haematobium infection is specifically linked to squamous cell carcinoma of the bladder (14). However, studies on the use of analgesics, water intake, coffee, and alcohol consumption have yielded mixed findings (11, 14, 15, 16, 17, 18, 19).

Due to recent shifts in geographic disparities in BC burdens and its strong correlation with modifiable risk factors, it is essential to understand country-specific BC risk factors for tailored public health interventions. (20) However, such studies have not yet been conducted in Ethiopia. Therefore, the objective of this study was to examine the association of occupational exposure to chemical substances and bladder cancer among patients admitted to selected hospitals in Ethiopia from May 01 to August 31 2023.

## METHODS

### Study Setting and Design

A matched case-control study was conducted between May 01 and August 31 2023 at four hospitals in Addis Ababa, Ethiopia—Tikur Anbessa Specialized Hospital, St. Paul’s Hospital Millennium Medical College, Yekatit 12 Hospital Medical College, and Menelik II Referral Hospital. These hospitals were chosen using a random method from the 14 hospitals with urology departments in the setting.

### Population and Eligibility

The source population comprised all adult patients admitted to either urology(cases) or non-urology departments (controls) in selected hospitals between May and August 2023. The study population for both cases and controls were determined by meeting the eligibility criteria. Eligible cases were patients diagnosed with histo-pathologically confirmed primary BC, while controls were patients meeting criteria of no history of bladder cancer or any other cancer, and absence of lower urinary tract symptoms. Age (±5 years), sex, and smoking status were matched in 68% of the participants.

### Sample Size Determination and Sampling Technique

The sample size was calculated using the formula for case control study with categorical outcome, assuming a 95% confidence level, 80% power and a case-to-control ratio of 1:2.5. Proportions of exposure to high-risk jobs (textile, metal, and chemical industries) were derived from a previous study (P2 = 0.35, P1 = 0.145) (9). Additionally, a 10% non-response rate and a design effect of 1.5 were considered, resulting in a final sample size requirement of 323 participants, comprising 92 cases and 231 controls.

For every two cases, five controls were randomly selected. Finally, a total of 90 cases and 213 controls: Tikur Anbessa Specialized Hospital (50cases and 123controls), St. Paul’s Hospital Millennium Medical College (18cases and 46 controls), Yekatit 12 Hospital Medical College (10cases and 20controls), and Menelik II Referral Hospital (12 cases and 24controls) were included in the study.

### Operational Definitions

Occupational exposure was classified as follows:

- High-exposure occupations were characterized by exposure to substances like aromatic amines, amino-biphenyl, PAHs, and azo dyes. They included dye manufacturing, leather industry, painting, rubber industry, mechanics, printing industry, textile industry, metalworking, truck driving, aluminum industry, hairdressing, and transportation equipment industry (21, 22).
- Low-exposure occupations were those which do not typically involve exposure to known or likely bladder carcinogens. They included clerical jobs (office jobs), construction workers, salesmanship, farming, and housekeeping. Employment for less than one year was also considered to pose low occupational risk across all types of occupations, hence categorized as low risk (21, 22).
- Duration of employment was classified as high risk if greater than 10 years and low risk if less than 10 years (23).
- Smoking status was categorized as non-smoker for those with no history of smoking or smoked fewer than 100 cigarettes in their lifetime, ex-smoker for individuals who had quit smoking for at least the past year, and smoker for current smokers (24).
- Water intake was considered low risk if greater than 1400 ml per day and high risk if less than 400 ml per day (15).
- Alcohol intake was categorized as heavy consumption for those consuming three or more drinks per day, and moderate consumption for those consuming fewer than three drinks per day (17).
- Analgesic use was defined as regular if used at least four times a week over the last month, and non-regular if used less frequently (18).

### Data Collection Procedures and Quality Assurance

A pre-tested electronic questionnaire was developed based on relevant literature to align with the study objectives, covering socio-demographic characteristics, occupational risks, and non-occupational risks. Three BSc nurses from selected hospitals who were assigned for data collection underwent a one-day training session. Data collection began with BC patients. The collected data was categorized by age, sex, and smoking status to align with control group variables. Data consistency and completeness were verified before coding and analysis commenced.

### Statistical Analysis

The extracted data underwent cleaning and analysis using SPSS version 26 software. Categorical covariates were summarized with frequencies and percentages while median with interquartile range was used for the numerical variable. Differences in underlying characteristics between cases and controls were assessed using the Chi-square test and Mann-Whitney U test with statistical significance set at p ≤ 0.05. Assumptions for the Chi-square test were verified prior to analysis to ensure data adherence to criteria. Furthermore, as the numerical variable did not meet the normality assumption under the Kolmogorov-Smirnov and Shapiro-Wilk tests, the Mann-Whitney U test was utilized.

To examine the association of occupational exposure to chemicals and BC, a multivariable binary logistic regression model was fitted. Variable selection was guided by univariate analysis at a significance level of 25% and by considering clinical relevance of the matched factors. The model generated adjusted odds ratios (AORs) with 95% CI, identifying variables with p ≤ 0.05 as statistically associated with bladder cancer. The model fitness was evaluated using the Hosmer-Lemeshow test, yielding a p-value of 0.152, indicating good fit as the test did not reject the hypothesis of adequate fit to the data.

## RESULTS

### Socio Demographic Characteristics

Among the 303 patients studied, 90(29.7%) were cases and 213 (70.3%) were controls with 98% and 92% response rate respectively. Age (±5 years), sex, and smoking status were matched in 68% (207) of the participants. The majority of the participants were age 60 years and above (62.2% of cases, 59.1% of controls), male (72% of cases, 73% of controls) and non-smokers (61.1% of cases, 64.8% of controls). Ex-smokers were 21.1% of cases and 13.6% of controls, while current smokers were 17.8% of cases and 21.1% of controls. A significant proportion of participants were from Addis Ababa (53.1%), Oromia (19.1%), and Amhara (13.9%), with the remainder from various other regions of Ethiopia. Most participants had at least a primary education (67%) and earned an average monthly income of less than 10,000 birr (94.1%).

Based on chi-square test result, a significantly higher proportion of bladder cancer patients had less than primary education (cannot read or write or can only read and write) compared to controls (61.1% vs. 21.1%, p<0.001). Otherwise, there was no significant difference in terms of other factors.

### Non-Occupational Exposures

Having a first-degree family history of BC (6.7% VS 0%), p=0.001), recurrent UTI (47.8% VS 8.9%, p<0.0001), pelvic radiation (15.6% VS 4.2%, p=0.001), schistosomiasis infection (4.4% VS 0.0%, p=0.007), and better water intake i.e., at least 1.4 liters per day (87.8% VS 54.9%, p<0.0001) were significantly higher among cases than controls.

In contrast, controls had a significantly higher proportion of individuals with a BMI of 25 and above (31% vs. 1.1%, p<0.0001) and a history of receiving chemotherapy (7.5% vs. 0.0%, p=0.003) compared to cases. A significantly higher proportion of cases consumed coffee (93.3% vs. 72.3%), with a frequency of 3 or more cups per day (50.0% vs. 14.6%) and a duration of more than 20 years (74.4% vs. 50.2%) compared to controls, all with a p-value of <0.0001.

A significantly high proportion of BC patients used analgesics regularly (>4 per week in the last month) and in a high amount (at least 10 tablets of painkiller per week) compared to controls (66.7% VS 37.6%, p<0.0001) and (55.6% VS 20.7%, p<0.0001) respectively. In addition, there is a significant difference in the proportion of type of analgesics taken by BC patients as compared to controls; opioids (31.4% VS 1.9%), NSAIDS (24.4% VS 22.1%), paracetamol (11.1% VS 10.3%) and aspirin (0.0% VS 3.3%) with a p value of <0.0001. On the other hand, all BC patients took analgesics for less than 8 years while a significant proportion of controls took painkillers for more than 8 years (0.0% VS 1.4%, p<0.0001).

A significantly higher proportion of cases had factory near their residence compared to controls (14.4% VS 4.2%, p=0.002). Additionally, the median (IQR) distance of factory from residence was 200.0 (115.0-325.0) meters with a statistically significant difference in the mean rank of the distance between BC patients and controls (51meters VS 103.47meters, p<0.0001).

### Occupational Exposures

The majority 255 (84.2%) of participants had low-exposure occupations, with 88.9% of cases and 82.2% of controls falling into this category. From the various types of occupations collected among participants, a statistically significant proportion of BC cases were farmers as compared to controls (10.0% vs 7.5%, p<0.0001). And a significant proportion of controls were employed in office jobs and as merchants compared to cases (20.2% vs 1.1%, p<0.0001, 14.6% vs 3.3%, p<0.0001). Regarding duration of employment, a significantly higher proportion of controls had held one job for more than 10 years compared to cases (21.6% vs. 3.3%, p<0.0001). Additionally, a significant proportion of BC cases reported never having a job lasting more than a year compared to controls (73.3% vs. 20.7%, p<0.0001).

**Table 1:**
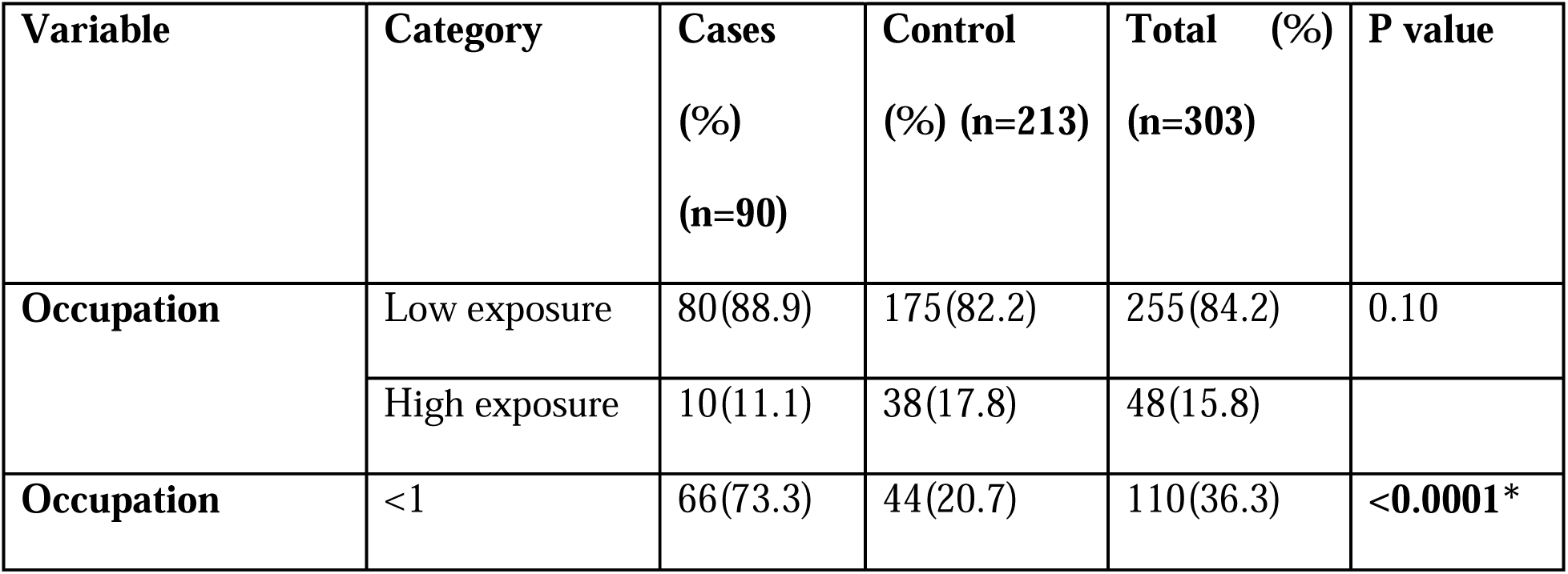
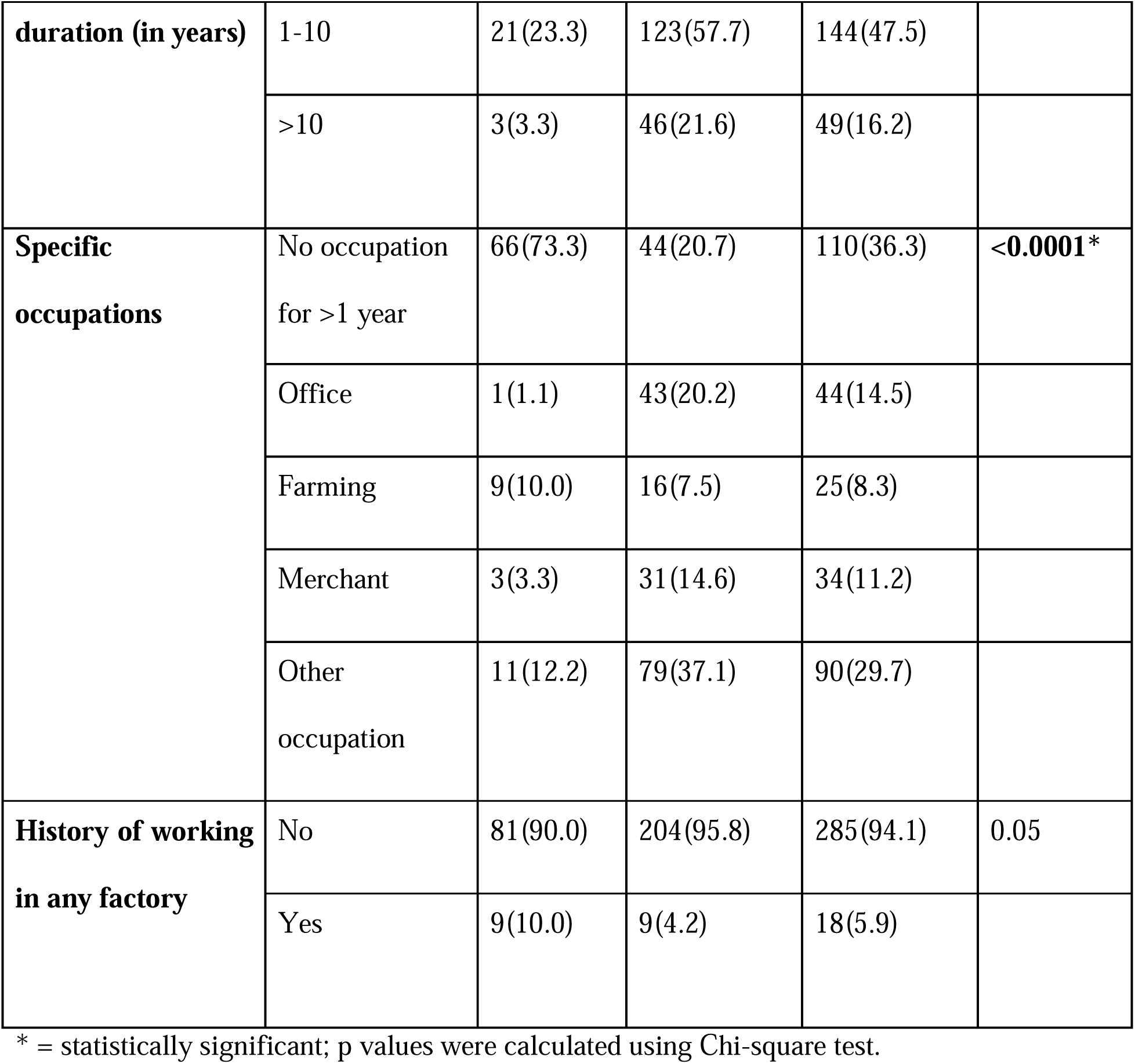
Occupational risk factors of bladder cancer, comparisons between bladder cancer patients and controls among patients in selected hospitals in Addis Ababa, Ethiopia, May 01 to August 31 2023 (n=303)

### Factors Associated with Bladder Cancer

After conducting univariate analysis at a 25% significance level, occupational risk, educational level, distance of factory from residence, source of drinking water, duration of coffee consumption, and regular analgesic use were found to be significantly associated with bladder cancer. In addition to these factors, clinical selection was done for age, sex, and smoking status to be included in the multivariable analysis. In the multivariable binary logistic regression, educational level, duration of coffee intake, and regular analgesic use were found to be significantly associated with bladder cancer at a 5% significance level. However, occupational risk did not show a significant association with bladder cancer (AOR:1.09; 95%CI= 0.44, 2.74; P=0.85).

**Table 2:** Socio-demographic characteristics of study participants among patients in selected hospitals in Addis Ababa, Ethiopia, May 01 to August 31 2023 (n=303)

| **Variable** | **Category** | **Cases (%)**<br>**(n=90)** | **Control (%)**<br>**(n=213)** | **Total (%)**<br>**(n=303)** | **P value** |
| --- | --- | --- | --- | --- | --- |
| **Age (in years)** | ≤49 | 14(15.6) | 21 (9.9) | 35(11.6) | 0.10 |
|  | 50-59 | 20 (22.2) | 66(31.0) | 86(28.4) |  |
|  | 60-69 | 26(28.9) | 75(35.2) | 101(33.3) |  |
|  | ≥70 | 30(33.3) | 51(23.9) | 81(26.7) |  |
| **Sex** | Female | 25(27.8) | 56(26.3) | 81(26.7) | 0.45 |
|  | Male | 65(72.2) | 157(73.7) | 222(73.3) |  |
| **Monthly income (in ETB)** | ≤4999 | 30(33.3) | 93(43.7) | 123(40.6) | 0.07 |
|  | 5000-9999 | 57(63.3) | 105(49.3) | 162(53.5) |  |
|  | ≥10,000 | 3(3.3) | 15(7.0) | 18(5.9) |  |
| **Education** | Cannot read or write | 18(20.0) | 17(8.0) | 35(11.6) | **<0.0001\*** |
|  | Can read and write | 37(41.1) | 28(13.1) | 65(21.5) |  |
|  | Primary education | 19(21.1) | 54(25.4) | 73(24.1) |  |
|  | Secondary education | 12(13.3) | 50(23.5) | 62(20.5) |  |
|  | Diploma and above | 4(4.4) | 64(30.0) | 68(22.4) |  |
| **Region** | Addis Ababa | 43(47.8) | 118(55.4) | 161(53.1) | 0.26 |
|  | Oromia | 18(20.0) | 40(18.8) | 58(19.1) |  |
|  | Amhara | 15(16.7) | 27(12.7) | 42(13.9) |  |
|  | Others | 14(15.5) | 28(13.1) | 42(13.8) |  |

ETB= Ethiopian birr; \* = statistically significant; p values were calculated using Chi-Square test.

**Table 2:**
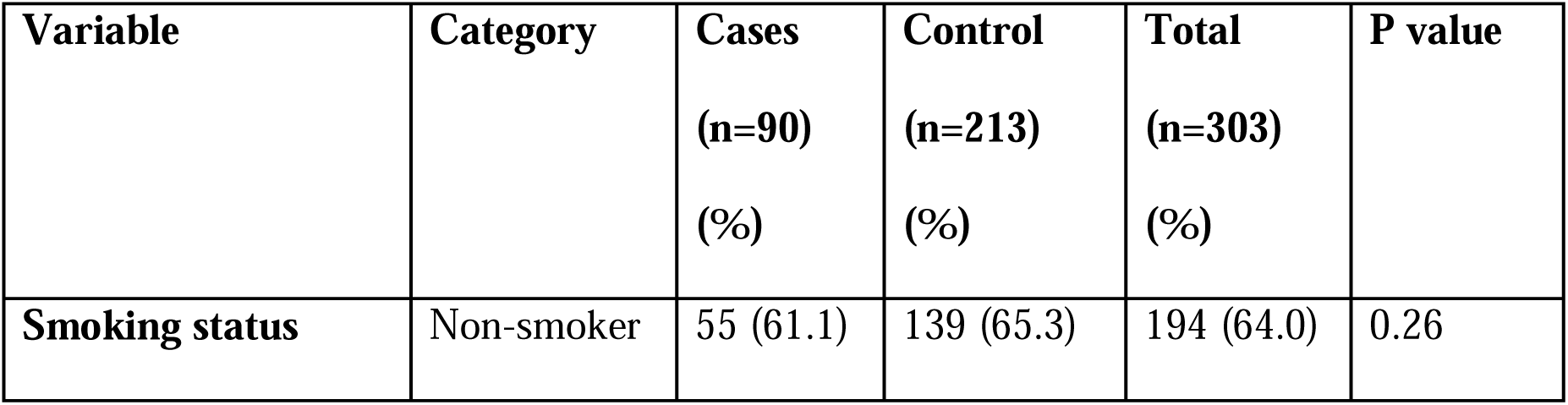
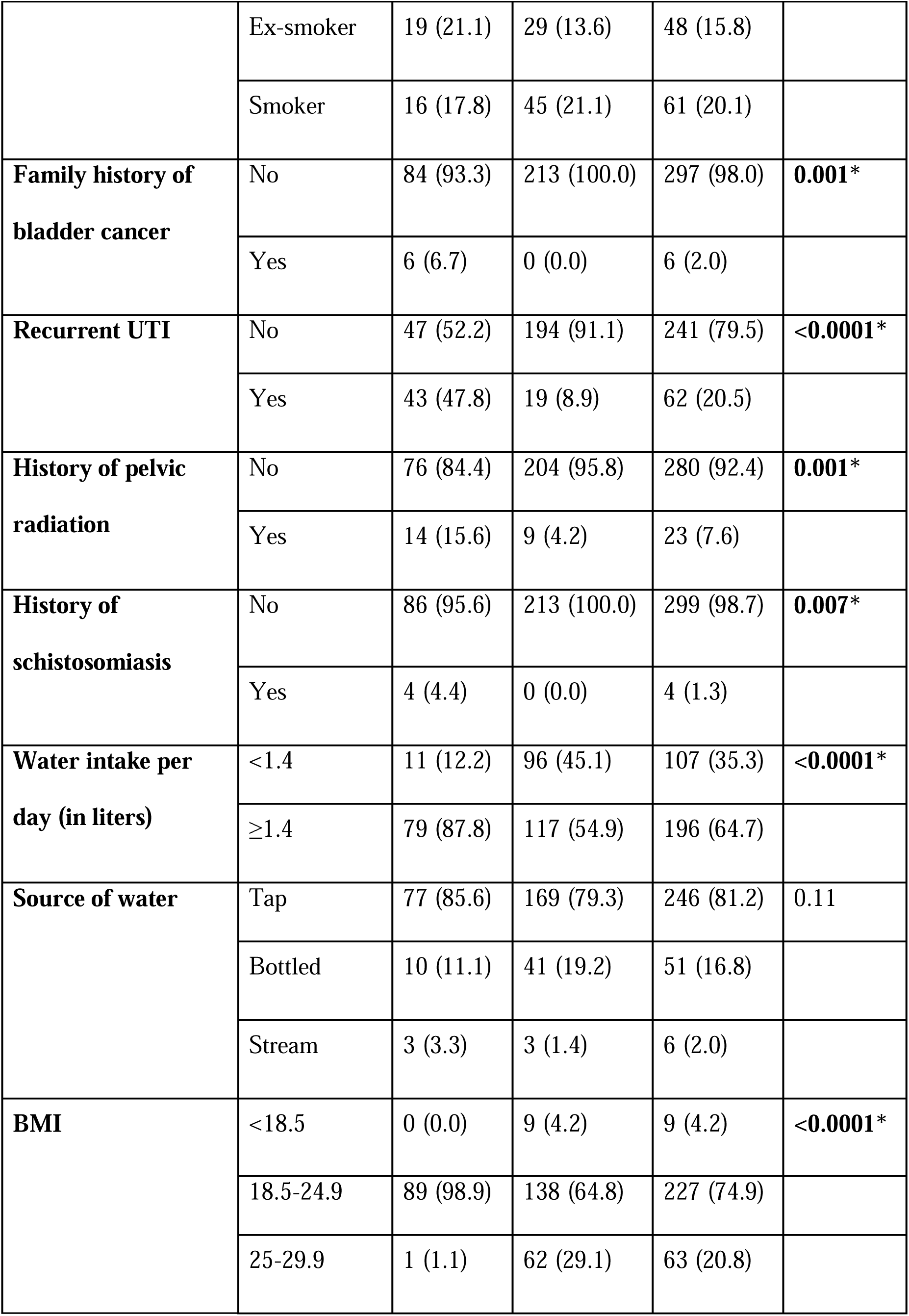
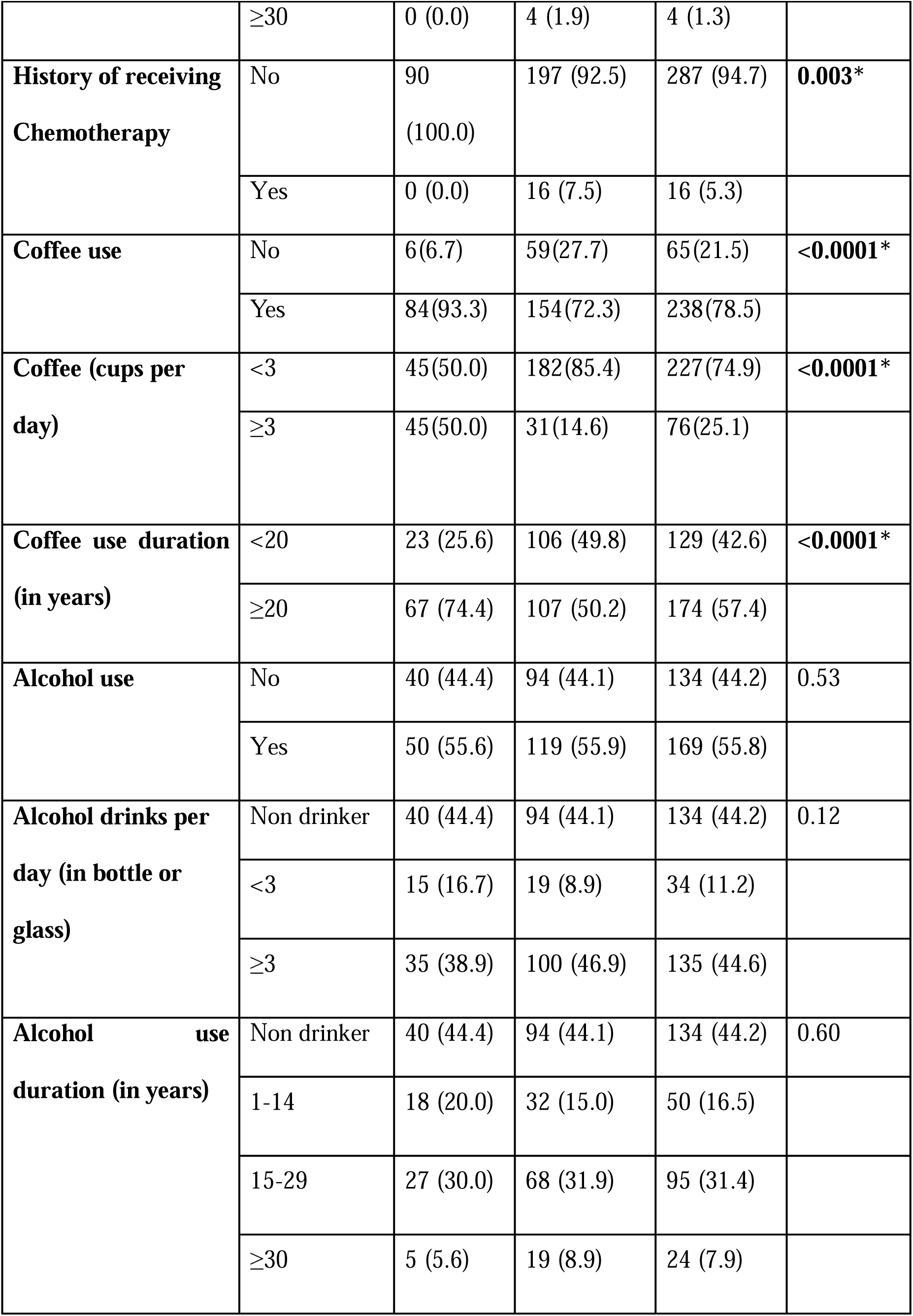
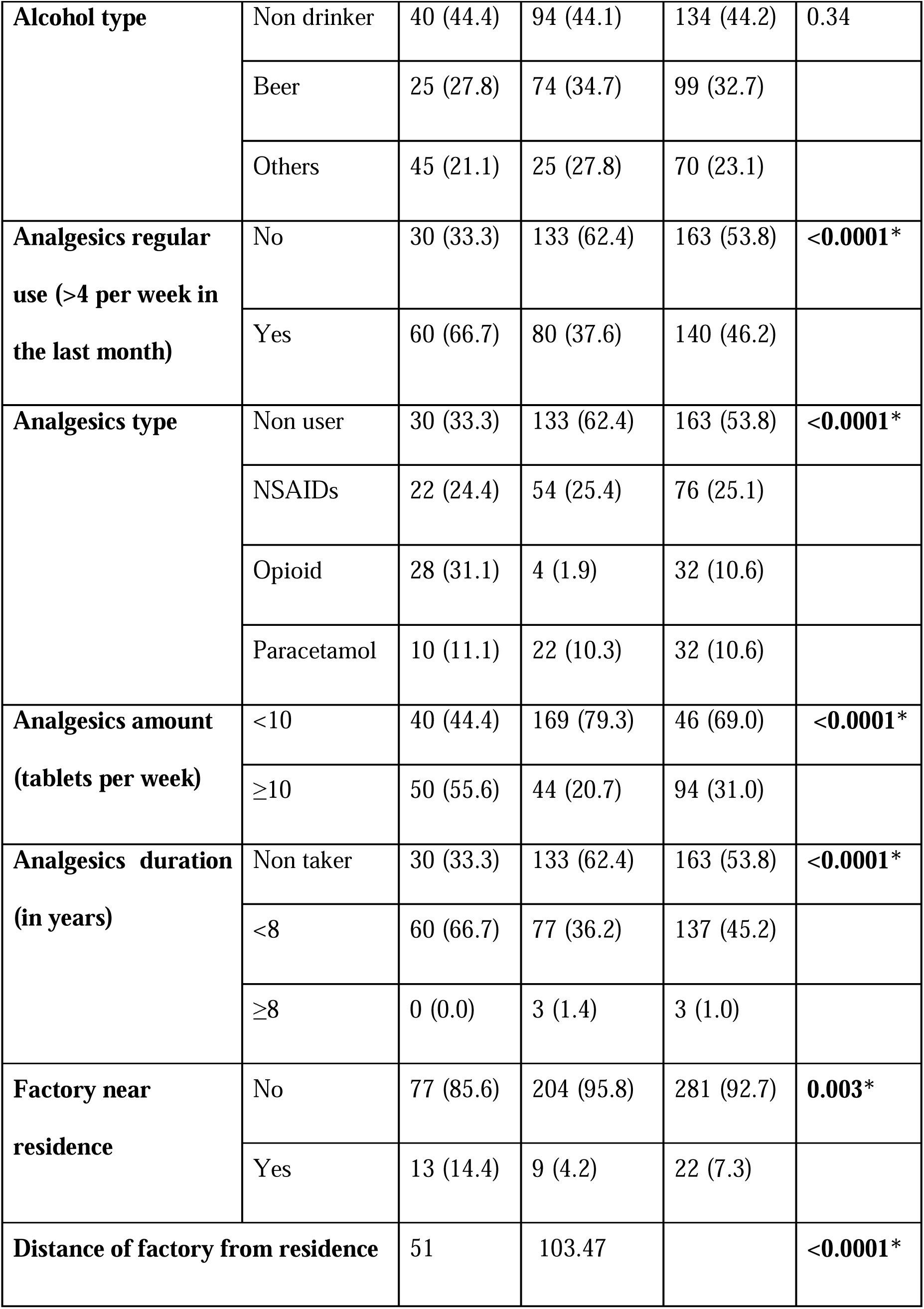
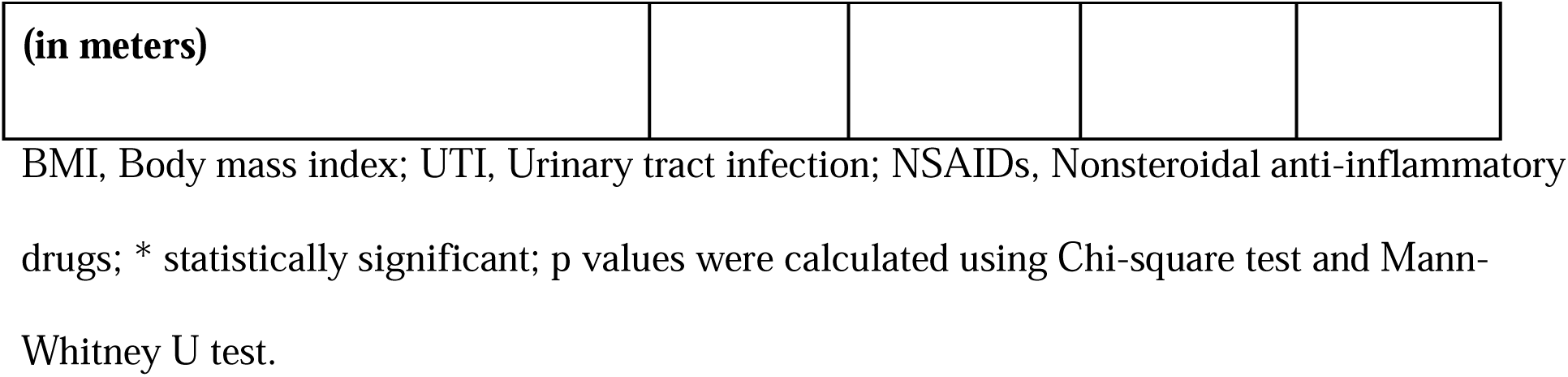
Non occupational risk factors of bladder cancer comparisons between bladder cancer patients and controls among patients in selected hospitals in Addis Ababa, Ethiopia, May 01 to August 31 2023(n=303)

| **Variable** | **Category** | **Cases**<br>**(n=90)**<br>**(%)** | **Control**<br>**(n=213)**<br>**(%)** | **Total**<br>**(n=303)**<br>**(%)** | **P value** |
| --- | --- | --- | --- | --- | --- |
| **Smoking status** | Non-smoker | 55 (61.1) | 139 (65.3) | 194 (64.0) | 0.26 |
|  | Ex-smoker | 19 (21.1) | 29 (13.6) | 48 (15.8) |  |
|  | Smoker | 16 (17.8) | 45 (21.1) | 61 (20.1) |  |
| **Family history of bladder cancer** | No | 84 (93.3) | 213 (100.0) | 297 (98.0) | **0.001\*** |
|  | Yes | 6 (6.7) | 0 (0.0) | 6 (2.0) |  |
| **Recurrent UTI** | No | 47 (52.2) | 194 (91.1) | 241 (79.5) | **<0.0001\*** |
|  | Yes | 43 (47.8) | 19 (8.9) | 62 (20.5) |  |
| **History of pelvic radiation** | No | 76 (84.4) | 204 (95.8) | 280 (92.4) | **0.001\*** |
|  | Yes | 14 (15.6) | 9 (4.2) | 23 (7.6) |  |
| **History of schistosomiasis** | No | 86 (95.6) | 213 (100.0) | 299 (98.7) | **0.007\*** |
|  | Yes | 4 (4.4) | 0 (0.0) | 4 (1.3) |  |
| **Water intake per day (in liters)** | <1.4 | 11 (12.2) | 96 (45.1) | 107 (35.3) | **<0.0001\*** |
|  | ≥1.4 | 79 (87.8) | 117 (54.9) | 196 (64.7) |  |
| **Source of water** | Tap | 77 (85.6) | 169 (79.3) | 246 (81.2) | 0.11 |
|  | Bottled | 10 (11.1) | 41 (19.2) | 51 (16.8) |  |
|  | Stream | 3 (3.3) | 3 (1.4) | 6 (2.0) |  |
| **BMI** | <18.5 | 0 (0.0) | 9 (4.2) | 9 (4.2) | **<0.0001\*** |
|  | 18.5-24.9 | 89 (98.9) | 138 (64.8) | 227 (74.9) |  |
|  | 25-29.9 | 1 (1.1) | 62 (29.1) | 63 (20.8) |  |
|  | ≥30 | 0 (0.0) | 4 (1.9) | 4 (1.3) |  |
| **History of receiving**<br>**Chemotherapy** | No | 90<br>(100.0) | 197 (92.5) | 287 (94.7) | **0.003\*** |
|  | Yes | 0 (0.0) | 16 (7.5) | 16 (5.3) |  |
| **Coffee use** | No | 6(6.7) | 59(27.7) | 65(21.5) | **<0.0001\*** |
|  | Yes | 84(93.3) | 154(72.3) | 238(78.5) |  |
| **Coffee (cups per**<br>**day)** | <3 | 45(50.0) | 182(85.4) | 227(74.9) | **<0.0001\*** |
|  | ≥3 | 45(50.0) | 31(14.6) | 76(25.1) |  |
| **Coffee use duration**<br>**(in years)** | <20 | 23 (25.6) | 106 (49.8) | 129 (42.6) | **<0.0001\*** |
|  | ≥20 | 67 (74.4) | 107 (50.2) | 174 (57.4) |  |
| **Alcohol use** | No | 40 (44.4) | 94 (44.1) | 134 (44.2) | 0.53 |
|  | Yes | 50 (55.6) | 119 (55.9) | 169 (55.8) |  |
| **Alcohol drinks per**<br>**day (in bottle or**<br>**glass)** | Non drinker | 40 (44.4) | 94 (44.1) | 134 (44.2) | 0.12 |
|  | <3 | 15 (16.7) | 19 (8.9) | 34 (11.2) |  |
|  | ≥3 | 35 (38.9) | 100 (46.9) | 135 (44.6) |  |
| **Alcohol use**<br>**duration (in years)** | Non drinker | 40 (44.4) | 94 (44.1) | 134 (44.2) | 0.60 |
|  | 1-14 | 18 (20.0) | 32 (15.0) | 50 (16.5) |  |
|  | 15-29 | 27 (30.0) | 68 (31.9) | 95 (31.4) |  |
|  | ≥30 | 5 (5.6) | 19 (8.9) | 24 (7.9) |  |
| **Alcohol type** | Non drinker | 40 (44.4) | 94 (44.1) | 134 (44.2) | 0.34 |
|  | Beer | 25 (27.8) | 74 (34.7) | 99 (32.7) |  |
|  | Others | 45 (21.1) | 25 (27.8) | 70 (23.1) |  |
| **Analgesics regular use (>4 per week in the last month)** | No | 30 (33.3) | 133 (62.4) | 163 (53.8) | **<0.0001\*** |
|  | Yes | 60 (66.7) | 80 (37.6) | 140 (46.2) |  |
| **Analgesics type** | Non user | 30 (33.3) | 133 (62.4) | 163 (53.8) | **<0.0001\*** |
|  | NSAIDs | 22 (24.4) | 54 (25.4) | 76 (25.1) |  |
|  | Opioid | 28 (31.1) | 4 (1.9) | 32 (10.6) |  |
|  | Paracetamol | 10 (11.1) | 22 (10.3) | 32 (10.6) |  |
| **Analgesics amount (tablets per week)** | <10 | 40 (44.4) | 169 (79.3) | 46 (69.0) | **<0.0001\*** |
|  | ≥10 | 50 (55.6) | 44 (20.7) | 94 (31.0) |  |
| **Analgesics duration (in years)** | Non taker | 30 (33.3) | 133 (62.4) | 163 (53.8) | **<0.0001\*** |
|  | <8 | 60 (66.7) | 77 (36.2) | 137 (45.2) |  |
|  | ≥8 | 0 (0.0) | 3 (1.4) | 3 (1.0) |  |
| **Factory near residence** | No | 77 (85.6) | 204 (95.8) | 281 (92.7) | **0.003\*** |
|  | Yes | 13 (14.4) | 9 (4.2) | 22 (7.3) |  |
| **Distance of factory from residence** |  | 51 | 103.47 |  | **<0.0001\*** |

|  |
| --- |
| (in meters) |

BMI, Body mass index; UTI, Urinary tract infection; NSAIDs, Nonsteroidal anti-inflammatory drugs; \* statistically significant; p values were calculated using Chi-square test and Mann-Whitney U test.

The odds of developing BC were 88.2% lower among people who have at least a primary level of education as compared to people who cannot read or write. (AOR=0.12, 95%CI= 0.05, 0.30, P<0.0001). The odds of developing bladder cancer were 2.36 times higher among individuals who consumed coffee for more than 20 years compared to those who consumed less or did not drink coffee (AOR=2.36, 95% CI=1.22, 4.56, p=0.01). Regular analgesic use (at least four times a week in the past month) was associated with 6.60 times higher odds of developing bladder cancer compared to non-regular users (<4 times a week in the past month) (AOR=6.60, 95% CI=3.34,13.02, p<0.0001).

**Table 3:**
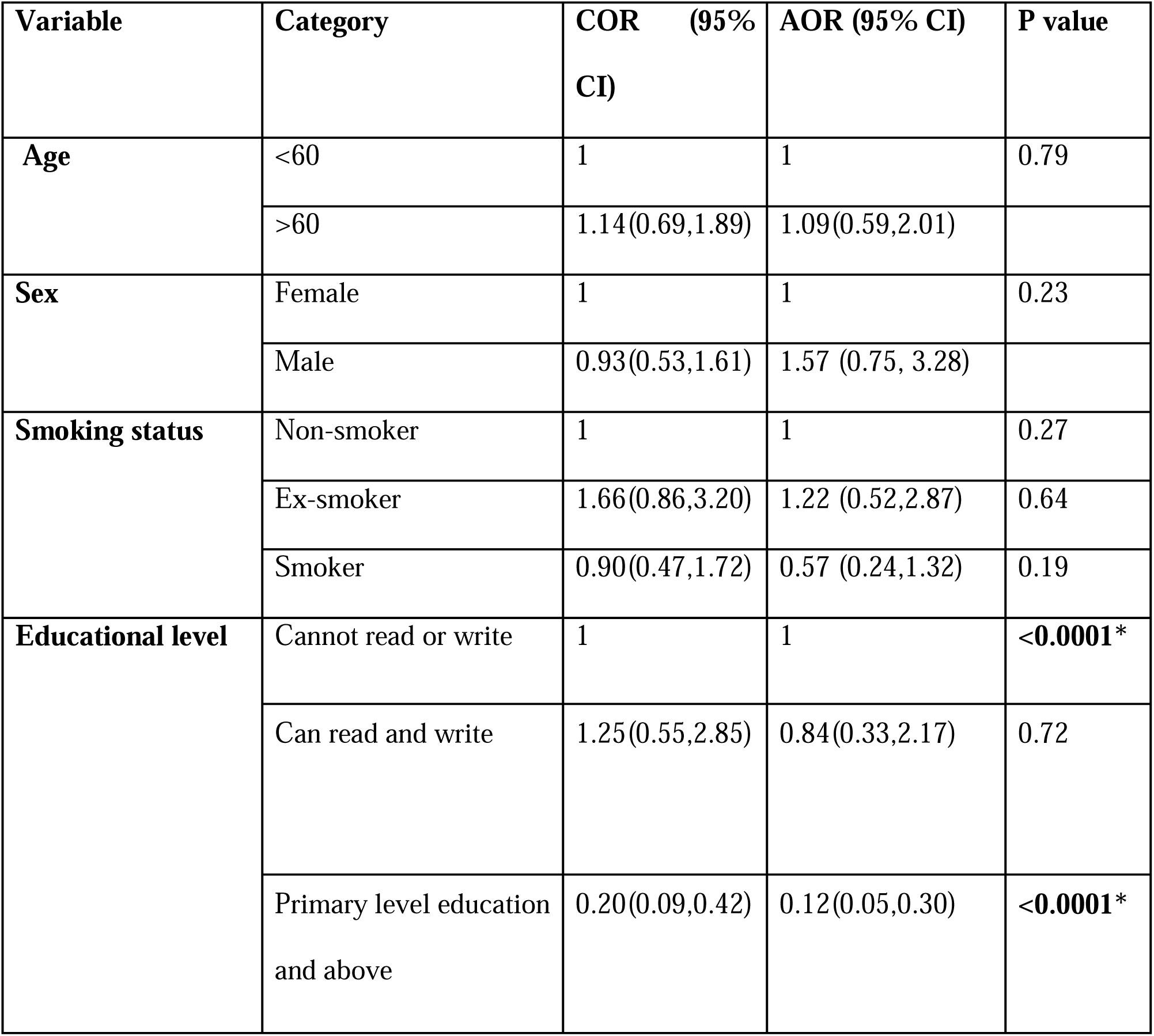
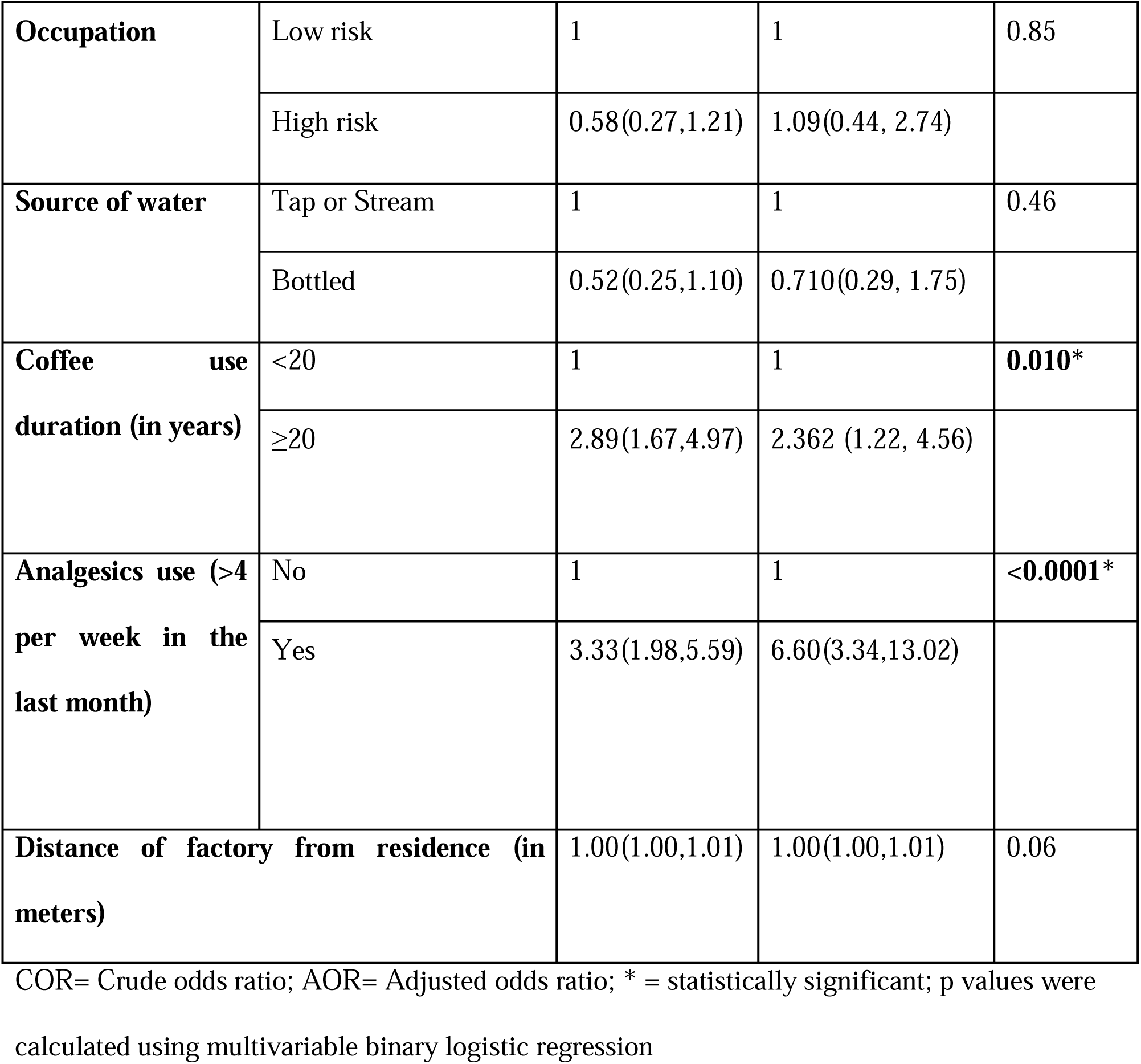
Results for the final multivariable binary logistic regression model among patients in selected hospitals in Addis Ababa, Ethiopia, May to August 2023 (n=303)

| Variable | Category | COR (95% CI) | AOR (95% CI) | P value |
| --- | --- | --- | --- | --- |
| Age | <60 | 1 | 1 | 0.79 |
|  | >60 | 1.14(0.69,1.89) | 1.09(0.59,2.01) |  |
| Sex | Female | 1 | 1 | 0.23 |
|  | Male | 0.93(0.53,1.61) | 1.57 (0.75, 3.28) |  |
| Smoking status | Non-smoker | 1 | 1 | 0.27 |
|  | Ex-smoker | 1.66(0.86,3.20) | 1.22 (0.52,2.87) | 0.64 |
|  | Smoker | 0.90(0.47,1.72) | 0.57 (0.24,1.32) | 0.19 |
| Educational level | Cannot read or write | 1 | 1 | <0.0001* |
|  | Can read and write | 1.25(0.55,2.85) | 0.84(0.33,2.17) | 0.72 |
|  | Primary level education and above | 0.20(0.09,0.42) | 0.12(0.05,0.30) | <0.0001* |
| **Occupation** | Low risk | 1 | 1 | 0.85 |
|  | High risk | 0.58(0.27,1.21) | 1.09(0.44, 2.74) |  |
| **Source of water** | Tap or Stream | 1 | 1 | 0.46 |
|  | Bottled | 0.52(0.25,1.10) | 0.710(0.29, 1.75) |  |
| **Coffee use duration (in years)** | <20 | 1 | 1 | **0.010\*** |
|  | ≥20 | 2.89(1.67,4.97) | 2.362 (1.22, 4.56) |  |
| **Analgesics use (>4 per week in the last month)** | No | 1 | 1 | **<0.0001\*** |
|  | Yes | 3.33(1.98,5.59) | 6.60(3.34,13.02) |  |
| **Distance of factory from residence (in meters)** |  | 1.00(1.00,1.01) | 1.00(1.00,1.01) | 0.06 |

COR= Crude odds ratio; AOR= Adjusted odds ratio; \* = statistically significant; p values were calculated using multivariable binary logistic regression

## DISCUSSION

The study aimed to investigate the association between occupational exposure to chemical substances and bladder cancer in Ethiopia. Out of 303 patients included, 90 (29.7%) had bladder cancer while 213 (70.3%) did not. Age, sex, and smoking status were matched in 68% of participants. Cases, compared to controls, had significantly higher proportions of individuals with less than primary education, coffee consumption, regular analgesic use, better water intake, residence around factories, family history of bladder cancer, recurrent UTIs, pelvic radiation, and schistosomiasis infection. Conversely, controls showed a significantly higher proportion of individuals with BMI ≥ 25 and history of chemotherapy. Most participants, 255(84.2%) had low-exposure occupations (88.9% cases, 82.2% controls), with cases showing a significantly higher proportion of farmers, and controls more often employed in office jobs or as merchants. Significantly higher proportion of controls also had a single job lasting over 10 years, while a significant proportion of cases reported having a job lasting less than a year.

After multivariable binary logistic regression analysis, the study did not identify a significant association between occupational exposures and BC, despite occupational factors being established as a risk according to The International Agency for Research on Cancer (IARC) (25). Past research has indicated that occupations lasting 10 or more years in high-risk fields are strongly linked to BC development (26). However, in this study, 73% of the bladder cancer participants had never held a job lasting more than one year, potentially reducing their exposure to occupational carcinogens. The lack of observed association in this study may also be due to the case-control ratio not providing sufficient statistical power.

On the other hand, educational level, coffee consumption, and analgesic use were found to have a statistically significant association with BC. People with at least a primary level of education had 88.2% lower odds of developing BC as compared to people who cannot read or write. This could be attributed to educated individuals often choosing low-exposure occupations such as office jobs. Moreover, higher education may promote healthier lifestyle habits such as increased fluid intake, smoking cessation, and prompt treatment of conditions like UTIs or schistosomiasis.

The study revealed that individuals who consumed coffee for more than 20 years had a 2.36 times increased odds of developing bladder cancer. Coffee’s dual nature—both potentially carcinogenic and antioxidant—has long been debated. Caffeine, a component of coffee, has been classified as potentially carcinogenic since the 1970s, and previous studies suggested a slight increase in bladder cancer risk associated with coffee consumption. This effect may be exacerbated by interactions with other environmental factors such as polycyclic aromatic hydrocarbons found in tobacco smoke, known to contribute to bladder cancer (27). From a clinical perspective, chemicals excreted through urine can directly affect the bladder wall before elimination, potentially leading to bladder cell dysplasia and cancer formation (1,28). However, a meta-analysis comprising studies from Europe, the USA, and Japan found no significant association between coffee consumption and bladder cancer (16). In this study, given Ethiopia’s significant cultural consumption of coffee, a long duration of coffee intake (>20 years) was reported by 57.4% of participants. This may suggest a potential geographic variation in the prevalence of long-term coffee consumption and hence its association with BC.

In this study, individuals who regularly used analgesics were found to have 6.60 times higher odds of developing bladder cancer. Given that study participants were hospitalized and predominantly over 60 years old, chronic pain and regular analgesic use were expected. This finding aligns with previous research indicating that analgesics containing phenacetin can be carcinogenic to the urinary tract, particularly with prolonged use exceeding 8 years (18). The literature on painkillers presents mixed findings regarding their impact on bladder cancer risk. A comprehensive meta-analysis incorporating 8 cohort studies and 9 case-control studies found no significant association between the use of paracetamol, aspirin, or other NSAIDs and bladder cancer risk (19). Similarly, this study did not identify a significant relationship between the use of various analgesics and BC, suggesting that the regular use of analgesics may be associated with BC rather than the specific type of drug used.

The study’s key strengths were the use of matching techniques and controlling a number of potential non-occupational factors. Although, the study was unable to assess specific chemical substances used in high exposure occupations due to recall bias. In addition, our study was conducted in hospital settings, which may not accurately reflect the conditions found in outpatient settings.

## CONCLUSIONS

The study did not find an association between bladder cancer and occupational exposure to chemicals. However, level of education, long-term coffee consumption, and regular analgesic use were identified as having a significant association with bladder cancer in the setting. Health care professionals are advised to educate patients about safe analgesic use. Furthermore, future multicenter case-control studies should be conducted to further explore the potential associations of these factors with bladder cancer.

## LIST OF ABBREVIATIONS

BC: Bladder cancer

IARC: International Agency for Research on Cancer

ILO: International Labor Organization

NSAIDS: Non-steroidal anti-inflammatory drugs

OHS: Occupational health services

PAHs: Polycyclic aromatic hydrocarbons

UTI: Urinary tract infection

## DECLARATION

### Ethics approval and consent to participate

The study was conducted after ethical clearance was obtained from the ethical committee of the School of Public Health at GAMBY Medical and Business College, Black Lion Specialized Hospital, St. Paul Hospital, and Addis Ababa Health Bureau. The ethical committees approved the use of verbal consent. Data collectors read and explained the informed consent to participants and documented consent on the Google Form sheet. The study posed no risks or negative consequences to the participants. Medical record numbers were used for data collection, and personal identifiers were not included in the research report to ensure confidentiality. Access to collected information was restricted to the principal investigator, and confidentiality was strictly maintained throughout the project.

### Consent for publication

Not applicable

### Availability of data and materials

The dataset used during the current study is available from the corresponding author on reasonable request.

## Authors’ Contribution

EAK conceived, designed, performed statistical analysis, and drafted the initial manuscript. TWL mentored and contributed to the design, statistical analysis, interpretation of findings and revision of the manuscript. All authors read and approved the final manuscript.

## Data Availability

All data produced in the present study are available from the corresponding author on reasonable request.

## Acknowledgment

The authors would like to thank GAMBY Medical and Business College for providing the necessary resources throughout the research endeavor. The authors would also like to thank everyone involved in the research through data collection, supervision, and facilitation.

